# Delayed introduction, contact variation, and susceptible dynamics explain spatial asynchrony during Korea’s large pertussis outbreak

**DOI:** 10.1101/2025.11.21.25340758

**Authors:** Sang Woo Park

**Affiliations:** School of Biological Sciences, Seoul National University, Seoul, Korea

## Abstract

Infectious disease outbreaks typically exhibit a large-scale spatial synchrony, reflecting coupling through shared environmental forcing and human mobility. A few exceptions to this theoretical expectation have been reported, but the conditions under which synchrony breaks down remain poorly understood. South Korea’s 2024–2025 pertussis outbreak provides a striking example, during which epidemic trajectories diverged substantially throughout the country: notably, the decline in spatial synchrony with distance was more pronounced than that reported in many previous outbreaks. Integrating high-resolution surveillance data from 252 municipalities and a Bayesian transmission model, I show that heterogeneity in introduction timing and differences in local contact levels can drive such fine-scale asynchrony even when all locations share identical seasonal forcing for transmission rates. In particular, regional variation in baseline contact levels can drive differential levels of susceptible depletion, thus shaping the potential for multiple epidemic waves. This analysis offers a unique perspective on spatiotemporal epidemic dynamics that depart from classical epidemic theory.

## Introduction

Spatiotemporal variation in epidemic dynamics offers a powerful means for understanding factors that determine the timing and magnitude of epidemic waves across populations [1, 2, 3, 4]. Ecological theory predicts that spatial coupling—via shared environmental forcing and human mobility—can synchronize epidemic dynamics between neighboring locations, where epidemic waves propagate through space [5, 6, 7]. Consistent with this theory, large-scale spatial synchrony has been observed across diverse pathogens, including measles [7], dengue [8], influenza [9], RSV [4], and SARS-CoV-2 [10], highlighting the near-ubiquity of spatial coupling in shaping epidemic dynamics. However, an exception to this pattern has been observed in pre-vaccination pertussis epidemics, which exhibited substantially weaker spatial correlation than measles epidemics reflecting a stronger sensitivity to stochasticity and transients [11].

Pertussis is a vaccine preventable respiratory disease, caused by the bacterium *Bordetella pertussis* [12]. Following the introduction of DTwP (diphtheria, tetanus, and whole cell pertussis) vaccine in 1954 and DTaP (diphtheria, tetanus, and acellular pertussis) vaccine in 1989 in Korea, the number of pertussis cases in Korea decreased dramatically, with less than 10 cases reported in 1995 compared to *>* 10, 000 cases reported annually around 1955–1963 [13]. However, pertussis cases have resurged in Korea and many other countries despite high vaccine coverage, raising concerns about the possibility of vaccine waning [14, 15, 16] and pathogen evolution [17, 18, 19].

Between 2024 and 2025, an unprecedented large pertussis outbreak occurred in Korea, resulting in more than 50,000 cases by early 2025 [20]. This outbreak marked a dramatic increase in the disease compared to 292 cases reported in 2023 and 21 cases reported in 2022, despite high vaccine coverage (91.1% of identified 2024 cases had a known vaccination history) [21, 20]. A considerable spatiotemporal variation in disease burden has been observed during the outbreak, but we currently have limited understanding of the underlying spatiotemporal variation in epidemic dynamics [20]. Interestingly, this outbreak exhibited a pronounced spatial asynchrony, which deviates from theoretical expectations based on spatial coupling, raising questions about mechanisms driving this fine-scale spatiotemporal heterogeneity in pertussis epidemic (Figure 1).

**Figure 1:**
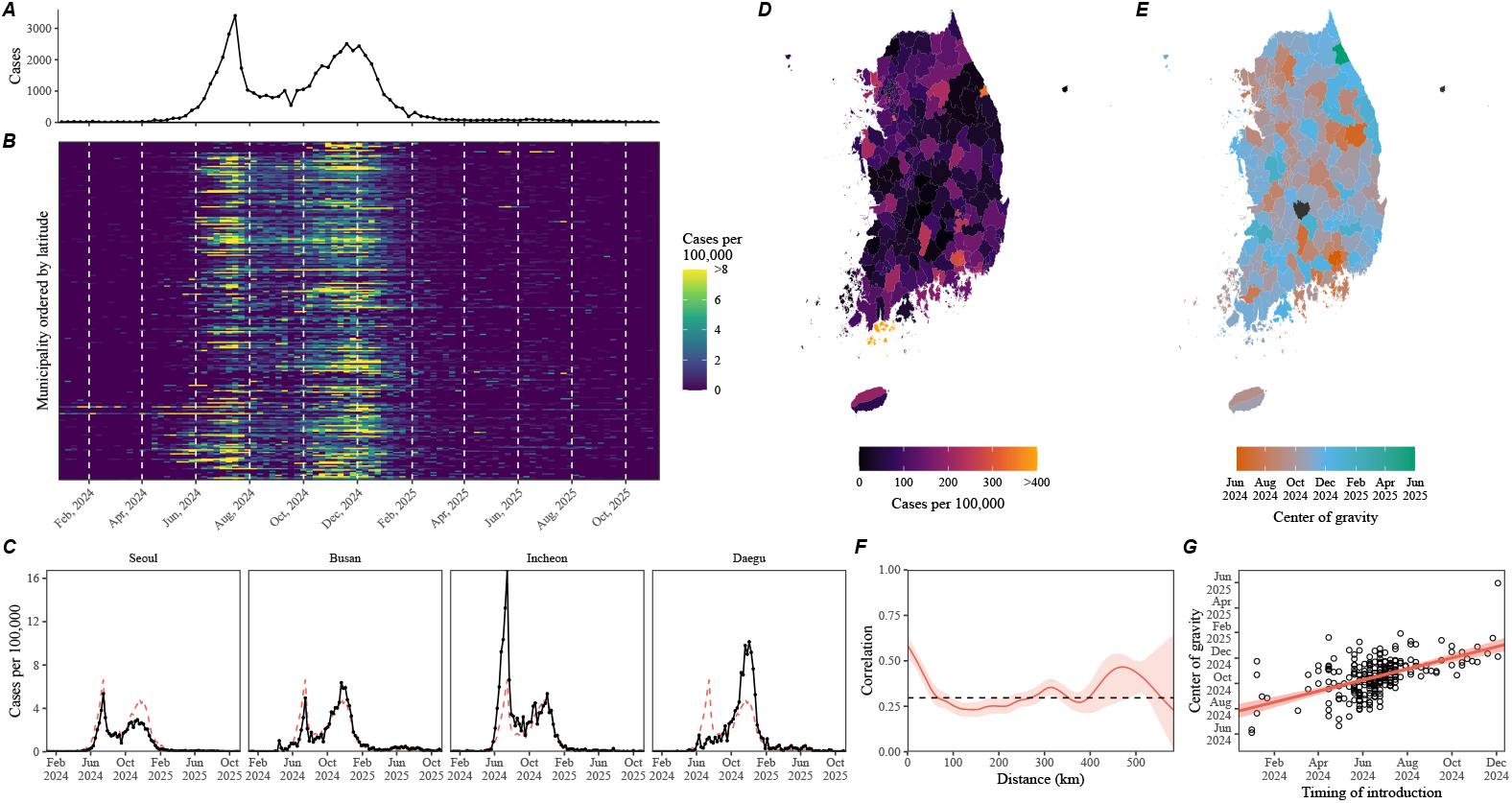
Spatiotempordal dynamics of pertussis outbreak in Korea, 2024—2025. (A) Total number of reported cases in Korea. (B) Number of reported cases per 100,000 individuals by municipality, ordered by latitude. (C) Number of reported cases per 100,000 individuals in four most populated cities in Korea. (D) Map of pertussis cases per 100,000 individuals between January 2024 and November 2025. (E) Map of center of gravity, which represents the mean timing of cases. (F) Spatial synchrony in logged cases as a function of distance. The red solid line and shaded regions represent the median estimate and the corresponding 95% confidence interval. The dashed horizontal line represents the mean correlation. (G) Relationship between center of gravity and the timing of introduction, defined as the first week when the number of reported cases is greater than 1 in 100,000. The red solid line and shaded regions represent the linear regression fit and the corresponding 95% confidence interval.

To understand mechanisms driving the apparent spatial asynchrony during the large pertussis outbreak in Korea, I combine a detailed spatiotemporal surveillance data from 252 municipalities with mathematical modeling approaches. I begin by showing that asynchrony in pertussis epidemics, captured by heterogeneous epidemic timing, is associated with differences in introduction delays. In contrast, the analysis of effective reproduction number reveals synchrony in underlying transmission patterns, suggesting that spatial asynchrony can arise despite synchrony in transmission. To test this hypothesis, I develop a spatiotemporal transmission model that allows for shared transmission term and variation in initial conditions and fit this model to observed data using Bayesian framework. Overall, the paper offers insight into how differences in initial conditions (i.e., introduction delays and initial susceptibility) can drive fine-scale spatiotemporal asynchrony in population dynamics.

## Materials and Methods

### Data

The weekly surveillance data on pertussis cases across 252 municipalities (1st week of 2024 to the 44th week of 2025) were obtained from a publicly available website, the Infectious Disease Portal, by the Korea Disease Control and Prevention Agency [22]. Population size data as of September 2025 were obtained from publicly available website by the Ministry of the Interior and Safety [23]. Vaccine coverage data were obtained from a publicly available website by the Korea Disease Control and Prevention Agency [24]. Longitude and latitude data for each municipality were obtained from a publicly available GitHub repository [25]. Finally, shape files used for constructing maps were obtained from a publicly available website [26].

### Center of gravity

I quantified the center of gravity to characterize variation in the mean timing of epidemic. Typically, the center of gravity is calculated based on annual incidence for recurrent epidemics. Since the paper focuses on analyzing a single outbreak, I used the entire time series of reported cases *C*_*t*_ to computer the center of gravity for each municipality:

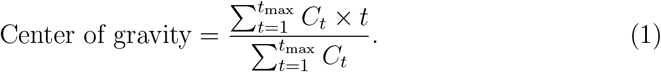

I then calculated the correlation coefficient between the center of gravity and the timing of introduction, defined as the first week when the number of reported cases is greater than 1 in 100,000.

### Spatial synchrony of reported cases

I characterized the spatial synchrony of pertussis epidemic, which captures changes in pairwise correlation as a function of distance [7]. I used logged values of reported cases between May 2024 and March 2025, excluding municipalities that had no reported cases (*n* = 250). Spatial synchrony was calculated using the ncf package in R [27]. I used 1,000 bootstraps to generate the median estimate and the corresponding 95% confidence intervals.

### Effective reproduction number

I calculated the effective reproduction number ℛ(*t*) to characterize changes in pertussis transmission over time. First, I took the case time series *C*_*t*_ and fitted a generalized additive model assuming a Poisson error to smooth the time series [28]. Then, I estimated ℛ(*t*) using the method of [29]:

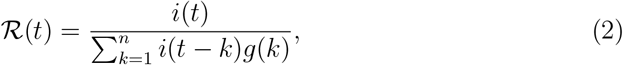

where *i*(*t*) is the smoothed incidence and *n* is the maximum length of generation interval in weeks (assumed to be 5 weeks). The generation-interval distribution *g*(*k*) was assumed to follow a gamma distribution with a mean of 9.47 days and a standard deviation of 6.22 days based on the estimated serial-interval distribution during this outbreak [30]. The reproduction number estimates can be unrealistically high when the number of infections is close to zero. Therefore, I only used estimates between May 2024 and March 2025 throughout the paper. I truncated all ℛ(*t*) estimates exceeding 5 to a maximum value of 5.

First, I compared correlation coefficients between ℛ(*t*) estimates and those between logged cases across all pairwise municipality combinations (Figure 2B in the main text). For this analysis, I only used data from municipalities with more than 400 total cases. Then, I compared the spatial synchrony for ℛ(*t*) estimates and synchrony for logged cases (Figure 2C in the main text). For this analysis, I used ℛ(*t*) estimates from all municipalities without any exclusion.

**Figure 2:**
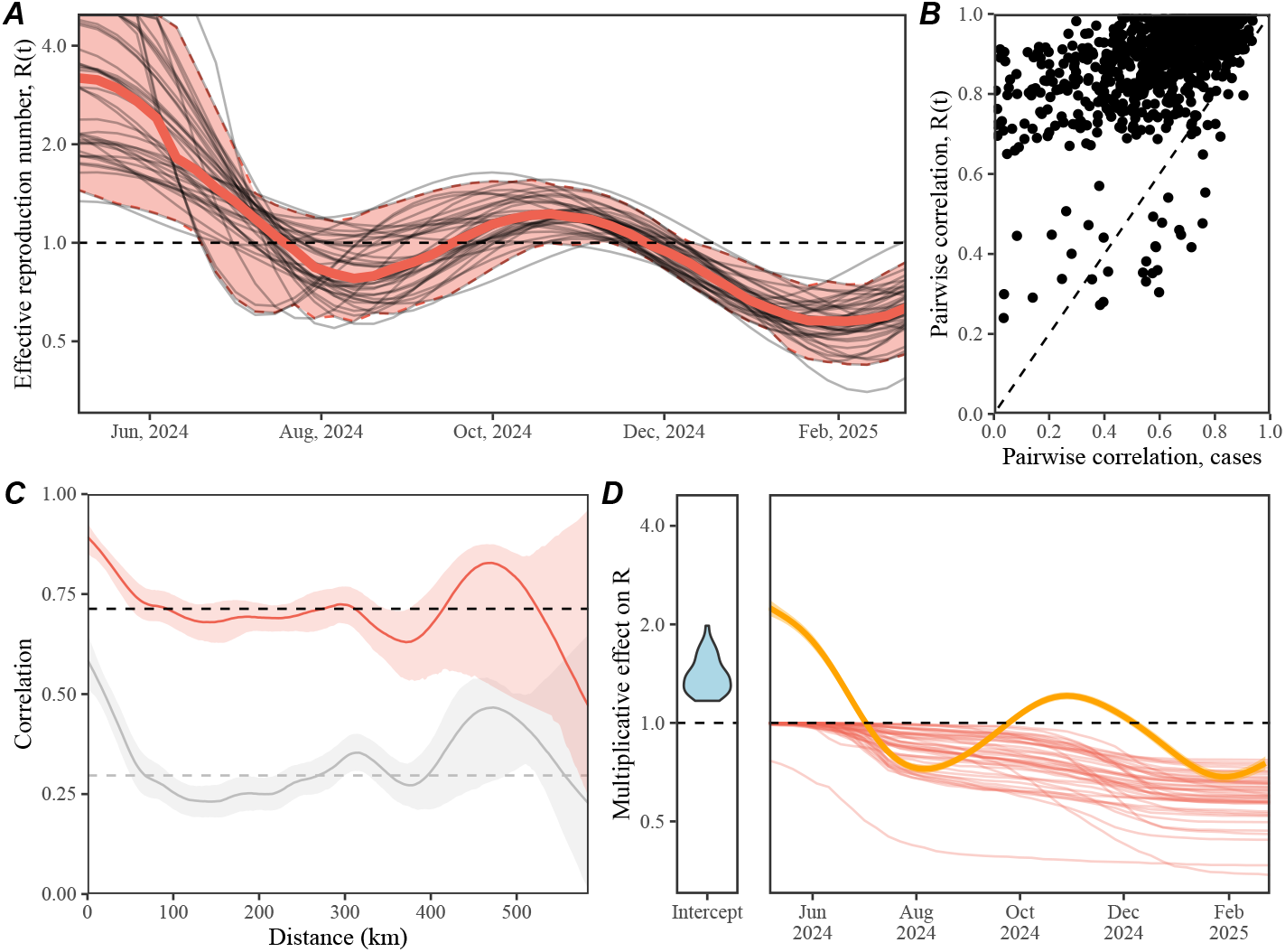
Spatial variation in the estimates of effective reproduction number. (A) Each black line represents the estimated effective reproduction number for a given municipality with more than 400 total cases. The red solid line and shaded regions represent the median and 95% quantiles of effective reproduction number estimates. (B) Relationship between correlation coefficient for ℛ(*t*) and correlation coefficient for reported cases across all pairwise municipality combinations with more than 400 total cases. (C) Spatial synchrony in ℛ(*t*) (red) versus logged cases (black) as a function of distance. The solid line and shaded regions represent the median estimate and the corresponding 95% confidence interval. The dashed horizontal line represents the mean correlation. (D) Estimated multiplicative effects on ℛ(*t*) from the regression analysis. The blue shaded violin represents the distribution of intercept effect, where a separate intercept term is estimated for each municipality. The red lines represent the estimated susceptible depletion effect for each municipality. The orange line and shaded regions represent the estimated changes in transmission from a smooth term and the corresponding 95% confidence interval.

Finally, I fitted a generalized additive model to logged values of ℛ(*t*) to quantify the effect of susceptible depletion and temporal variation in intrinsic transmission [31, 28, 32]:

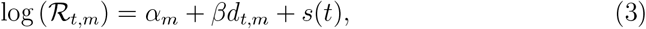

where ℛ_*t,m*_ represents ℛ(*t*) estimate at time *t* in municipality *m*; *α*_*m*_ represents the municipality-specific intercept term; *β* represents the effect of susceptible depletion; *d*_*t,m*_ represents the cumulative cases at time *t* in municipality *m*; and *s*(*t*) represents the smooth term capturing temporal variation in transmission. For this analysis, I only used ℛ(*t*) estimates from municipalities with more than 400 total cases.

### Transmission model

Finally, I fitted a simple transmission model using Bayesian inference to test whether variation in introduction timing and contact variation alone can explain the observed epidemic patterns. Specifically, I extended the SEIR model, which is commonly used for pertussis transmission [33], to allow for a joint estimation of a single, time-varying transmission term that is shared across all regions and a separate estimation of initial infected fraction, basic reproduction number, and reporting rates. To do so, I first discretized the SEIR model following the approach of [34]:

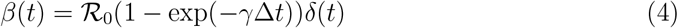

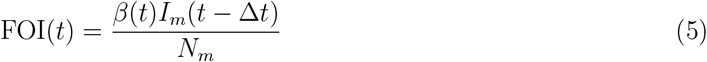

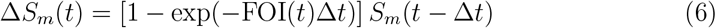

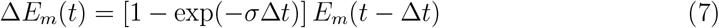

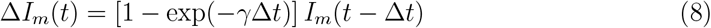

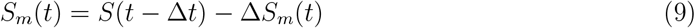

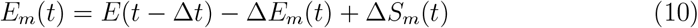

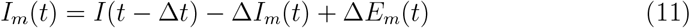

where *S*_*m*_(*t*), *E*_*m*_(*t*), and *I*_*m*_(*t*) represent the number of susceptible, exposed, and infectious individuals in municipality *m* at time *t*, respectively; *N*_*m*_ represents the population size of municipality *m*; Δ*t* represents the simulation time step, which assumed to be 1 week; *β*(*t*) represents the shared time-varying transmission rate; ℛ_0_ represents the shared basic reproduction number [35]; *δ*(*t*) represents the normalized time-varying transmission rate; *σ* represents the rate at which individuals develop infectiousness, which is assumed to be *σ* = −log(1−7*/*8) such that the mean latent period is 8 days; and *γ* represents the rate at which individuals recover, which is assumed to be *σ* = −log(1−7*/*15) such that the mean latent period is 15 days.

Here, I modeled changes in transmission using the *δ*(*t*) term, which is given a normal prior around 1:

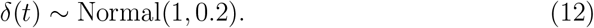

To allow for smooth variation in transmission, I also incorporated a random walk prior:

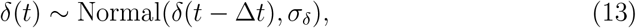

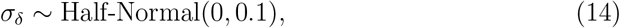

where a half-normal prior on *σ*_*δ*_ constrains the smoothness of *δ*(*t*). I also estimated a municipality-specific basic reproduction number:

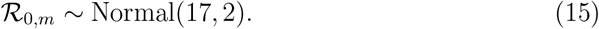

The municipality-specific observation process is modeled based on a Poisson distribution:

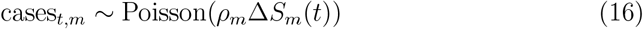

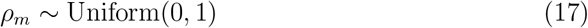

where cases_*t,m*_ represents the reported cases at time *t* in municipality *m*; *ρ*_*m*_ represents the municipality-specific reporting rate; and Δ*S*_*m*_(*t*) represents the expected number of new infections between time *t* − Δ*t* and *t*.

Finally, I estimated a municipality-specific initial infected fraction *i*_*m*_(0) and a shared initial susceptible fraction *s*(0) by imposing the following priors:

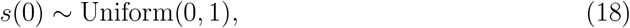

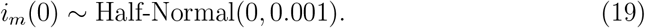

Then, the initial conditions for the model is specified as follows:

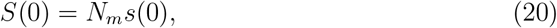

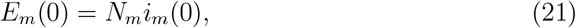

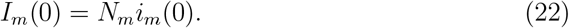

I simultaneously fitted this model time series data between May 2024 and March 2025 from all municipalities with more than 400 total cases (a total of 42 municipalities) using a Bayesian inference software rstan using 4 chains, each with 2000 iterations [36]. Convergence was assess by ensuring low R-hat (*<* 1.01), high effective sample size (*>* 800), no divergent transitions, and no iterations that exceeded the maximum tree depth.

To assess the model fit, I computed R squared values for each municipality. This is done by calculating the squared value of the correlation coefficient between logged values of the observed cases+1 and a posterior median of the logged predictions *ρ*_*m*_Δ*S*_*m*_(*t*).

Finally, I evaluated the impact of initial conditions on pertussis epidemic dynamics by varying ℛ_0_ between 14.2 and 22.2 and *i*(0) between 5 × 10^*−*6^ and 2.3 × 10^*−*3^. For each simulation, I quantified the center of gravity.

As an alternative hypothesis, I also considered a model assuming identical ℛ_0_ across municipality and estimated a municipality-specific initial susceptible fraction *s*_*m*_(0). Same prior distributions were used for this analysis.

### Transmission model validation

I validated the model by fitting the model to remaining municipalities (i.e., those with less than 400 total cases). In doing so, I assumed that changes in transmission rate *δ*(*t*) is known and only estimate the initial conditions, *s*(0) and *i*_*m*_(0), basic reproduction number ℛ_0,*m*_, and the reporting rate *ρ*_*m*_. For this validation, I used the optimization function in rstan [36] rather than performing a full Bayesian inference. Then, I computed R squared values for each municipality in the same way as before.

### Relationship between estimated initial susceptible and vaccine coverage

I assessed the relationship between estimated initial susceptible from the alternative model and vaccine coverage using linear regression. Specifically, assuming that the waning of vaccinal immunity is the main cause for pertussis re-emergence, I compared DTaP (Diphtheria-Tetanus-Pertussis) vaccine coverage for 3 years old as of 2015 and the estimated initial susceptible in each municipality and performed a linear regression. I used vaccine coverage as of 2015 because this was the oldest data that were publicly available. I considered using estimates from municipalities with *>* 400 cases, since the parameter estimates are expected to be more reliable.

## Results

### Asynchrony in pertussis epidemics associated with introduction delays

First, I begin by characterizing the spatiotemporal dynamics of the pertussis epidemic in Korea. The outbreak consisted of two waves that peaked around late July and December 2024 (Figure 1A) and exhibited large spatial heterogeneity in timing (Figure 1B,C) and magnitude (Figure 1D). For example, both Seoul, the capital city located in the northwest region, and Busan, the second most populated city located at the southeast end, experienced two waves, whereas Daegu, the fourth most populated city located *<* 100km away from Busan, only experienced the second wave (Figure 1C; see Figure S1 for epidemic dynamics in other regions).

To further quantify spatial heterogeneity in pertussis waves across Korea, I calculate the center of gravity (Figure 1E), which represents the mean timing of cases [1]. Notably, I find a sharp contrast between early (orange) and late (light blue) waves between many neighboring municipalities (Figure 1E), suggesting spatial asynchrony in pertussis dynamics. To systematically evaluate this apparent asynchrony, I estimate the spatial synchrony function, which captures changes in correlation in epidemic dynamics as a function of distance [7]. The spatial synchrony function confirms a sharp reduction in cross-correlation with distance (Figure 1F). For example, within 100km of distance, the cross-correlation drops below 0.25. By comparison, pertussis epidemics in the US maintained cross-correlations above 0.2 even at 1,000 km [37], highlighting the fine-scale asynchrony in pertussis epidemics in Korea. Interestingly, the variation in the center of gravity significantly correlates with the introduction timing (*ρ* = 0.53; 95%CI: 0.44—0.62; Figure 1G), suggesting that differences in introduction timing may contribute to the apparent spatial asynchrony.

### Synchrony in temporal changes in pertussis transmission

I then ask whether the observed epidemic asynchrony can be explained by the differences in transmission patterns. To do so, I quantify the effective reproduction number ℛ(*t*), which measures the average number of new infections caused by a single infected individual [38, 29]. For this analysis, I focus on municipalities with more than 400 total cases reported because ℛ(*t*) estimates can be unstable when the number of infections are low.

Notably, ℛ(*t*) estimates appear to be largely homogeneous across different regions, despite the apparent heterogeneity in the observed epidemic dynamics (Figure 2A). Comparisons of correlation coefficients in ℛ(*t*) versus correlation coefficients in reported cases between all pairwise municipality combinations confirm this observation (Figure 2B). In 830 out of 861 municipality combinations, correlation in ℛ(*t*) is higher than correlation in cases. This is reflected in the spatial synchrony function for ℛ(*t*) (Figure 2C), where the mean cross-correlation for ℛ(*t*) (*ρ* = 0.71) is considerably higher than that for cases (*ρ* = 0.30).

To better understand factors driving changes in ℛ(*t*), I perform a regression analysis using a generalized additive model with three covariates [31, 28, 32]: (1) a municipality-level fixed effect allowing each municipality to have its own intercept; (2) a cumulative-cases term capturing the effect of susceptible depletion; and (3) a single smooth term shared across municipalities, which allows for temporal changes in pertussis transmission over time, such as seasonal forcing. First, the smooth term suggests a strong transmission reduction around August and February, which coincide with the timing of school breaks (Figure 2D). Second, the model estimates a significant effect of effect of susceptible depletion (Figure 2D; *p <* 0.001). Notably, this simple regression model explains 84% of variance in ℛ(*t*), suggesting that the observed temporal fluctuations in transmission can be largely explained by school-related seasonality and the gradual depletion of susceptible individuals. Taken together, the analysis of center of gravity and ℛ(*t*) suggest that differences in introduction timing and susceptible host dynamics are likely main drivers for spatial asynchrony in pertussis epidemics in Korea, rather than variation in seasonal transmission patterns.

### Mathematical modeling of fine-scale spatio-temporal pertussis epidemic

To test whether variation in introduction timing and susceptible host dynamics can explain the observed epidemic patterns, I extend a standard susceptible-exposed-infected-removed (SEIR) model [33] to jointly estimate a single, time-varying transmission term, which is shared across all regions. In particular, the transmission rate *β*_*r*_(*t*) in region *r* is modeled as follows:

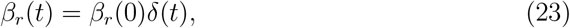

where *β*_*r*_(0) represents the baseline level transmission rates, which accounts for variation in baseline contact levels and is estimated separately for each region *r*. Then, *δ*(*t*) represents a time-varying transmission term, which are shared across all regions and captures a multiplicative change in transmission over time. I also allow the initial susceptible fraction *S*(0) to be shared across all regions to capture the average level of susceptibility. Initial infected fraction *I*_*r*_(0) and reporting rate *ρ*_*r*_ are allowed to vary across regions, where variation in *I*(0) effectively captures differences in introduction timing. For this analysis, I also focus on municipalities with more than 400 total cases. The model is fitted using a Bayesian inference software rstan [36].

This simple transmission model that assume shared transmission rate can capture fine-scale spatiotemporal variation of pertussis outbreak in Korea (Figure 3A) with a median R squared value of 0.86 (95% quantile: 0.71—0.92; Figure 3B). Furthermore, I find a near-perfect correlation between the predicted and observed center of gravity (Figure 3C). These results suggest that the model assuming shared time-varying transmission can sufficiently explain the fine-scale spatial asynchrony observed during the pertussis outbreak in Korea. So how do contact levels vary across regions and transmission change across time?

**Figure 3:**
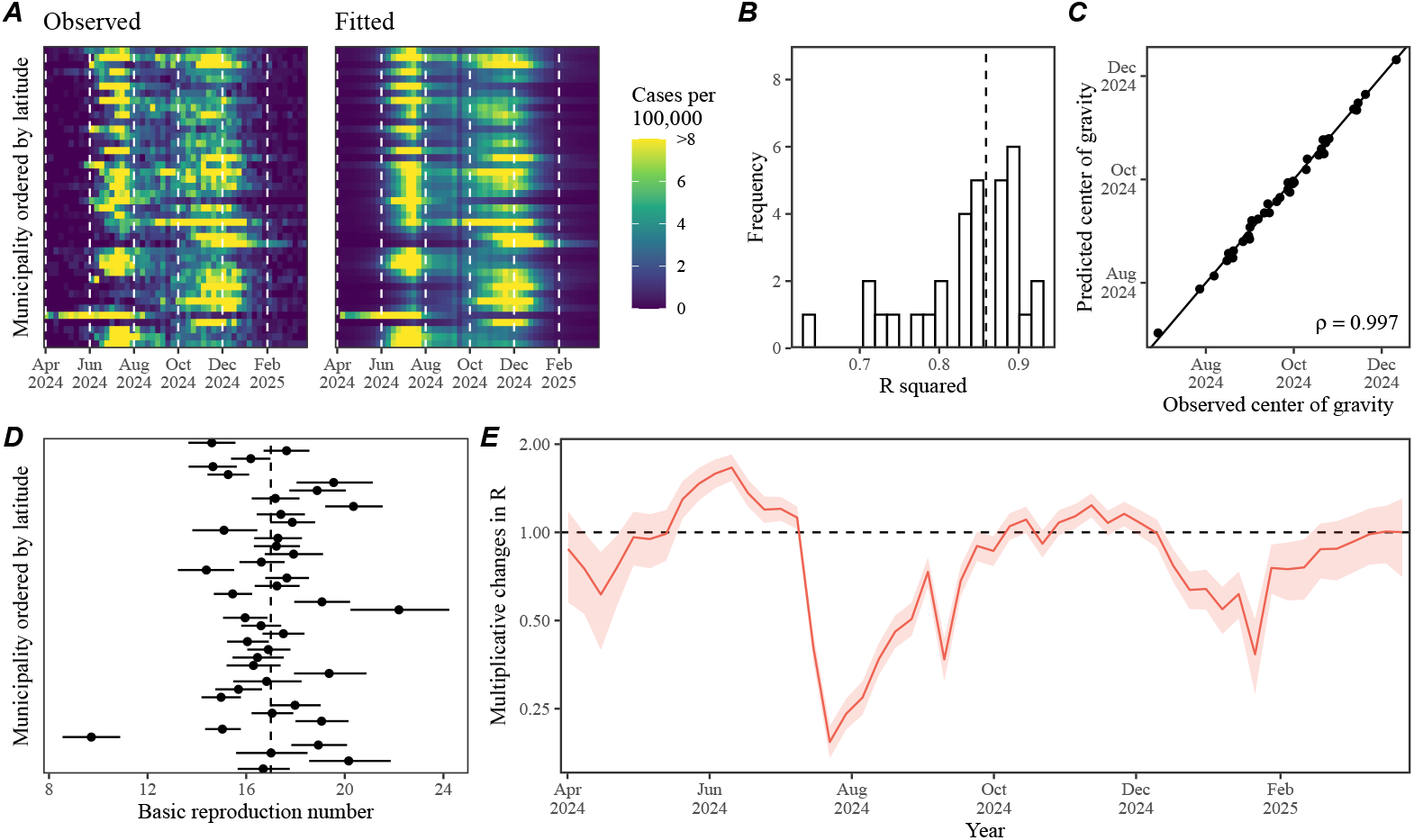
Fine-scale spatiotemporal variation in pertussis epidemic can be explained with a simple SEIR model. (A) Comparisons between the observed and predicted epidemic dynamics across municipalities with more than 400 total cases, ordered by latitude. (B) The bar plot represents the distribution of R squared values for model fits. The vertical dashed line represents the median. (C) Relationship between the estimated and predicted center of gravity. The solid line indicated the one-to-one relationship. (D) Estimated basic reproduction number ℛ_0_ across municipalities, ordered by latitude. Points and error bars represent posterior medians and corresponding 95% credible intervals. (E) Estimated time-varying pertussis transmission multiplier. The red line and shaded regions represent the posterior median and the corresponding 95% credible interval.

I estimate a considerable variation in ℛ_0_ across regions (Figure 3D). The median estimates for region-level ℛ_0_ vary from 9.7 to 22.1 with an average of 17, consistent with previous estimates [39, 16, 40]. The estimated initial susceptible fraction is 0.15 (95% CI: 0.14–0.16), which correspond to an initial effective reproduction number of 2.6 when combined with the average ℛ_0_ estimate—this is consistent with the initial ℛ (*t*) estimate. Finally, I also estimate a large reduction in transmission around late July 2024 and late January 2025 (Figure 3E). The estimated timing of transmission reduction is also consistent with results from the ℛ (*t*) analysis and coincides with the timing of school holidays in Korea. Parameter estimates for the initial infected fraction *I*(0) and reporting rates also reveal large variation across regions (Supplementary Figure S2). While estimates of ℛ_0_ or *I*(0) do not correlate with population sizes, reporting rate negatively correlates with population size (Supplementary Figure S3).

I validate the model by taking the estimated transmission term *δ*(*t*) and fitting the remaining parameters (i.e., initial infected, initial susceptible, basic reproduction number, and reporting probability) to data from the remaining 210 municipalities with less than 400 total cases (Figure S4). While model predictions appear consistent with the observed patterns (Figure S4A), I estimate a lower value for the median R squared: 0.59 (95% quantile: 0.01–0.85; Figure S4B). Nonetheless, this analysis suggests that the majority of asynchrony in pertussis epidemics can be explained via the differences in introduction timing and contact variation.

As an alternative hypothesis, I also considered the possibility that the spatiotemporal asynchrony may be driven by variation in susceptibility. To do so, I assumed a fixed basic reproduction number, ℛ_0_ = 17 and estimated a separate initial susceptible fraction *S*_*r*_(0) for each region and a shared time-varying transmission term *δ*(*t*). This alternative model provided nearly identical fits to data compared to the original model (Supplementary Figure S5–S6) and similar levels of out-of-sample validation (Supplementary Figure S7). However, I did not find any correlation between past vaccine coverage and the estimated levels of susceptibility (Supplementary Figure S7). Moreover, the estimated range for the initial susceptible fraction was unrealistically wide compared to the reported range for past vaccine coverage (Supplementary Figure S7). Thus, I tentatively conclude that a model assuming contact variation is likely more parsimonious and continue the analysis.

So how does variation in introduction timing and contact rates generate heterogeneity in epidemic waves? I address this question by simulating the model between April 2024 and April 2025 across plausible ranges of ℛ_0_ and *I*(0). First, increasing *I*(0) leads to an earlier epidemic because it permits earlier introduction of the infection to the population (Figure 4A). This is consistent with the earlier observations (Figure 1F). Similarly, increasing ℛ_0_ also results in an earlier epidemic. This is because high levels of transmission allows for a large first wave (Figure 4B), which prevents the second wave through the overcompensatory susceptible depletion (Figure 4C). In contrast, when ℛ_0_ is low, there will be enough susceptible hosts for the population to experience the second wave (Figure 4C). These results are consistent with an earlier finding, which showed that contact variation can explain a sudden shift in RSV seasonality from a later wave to an earlier wave [41].

**Figure 4:**
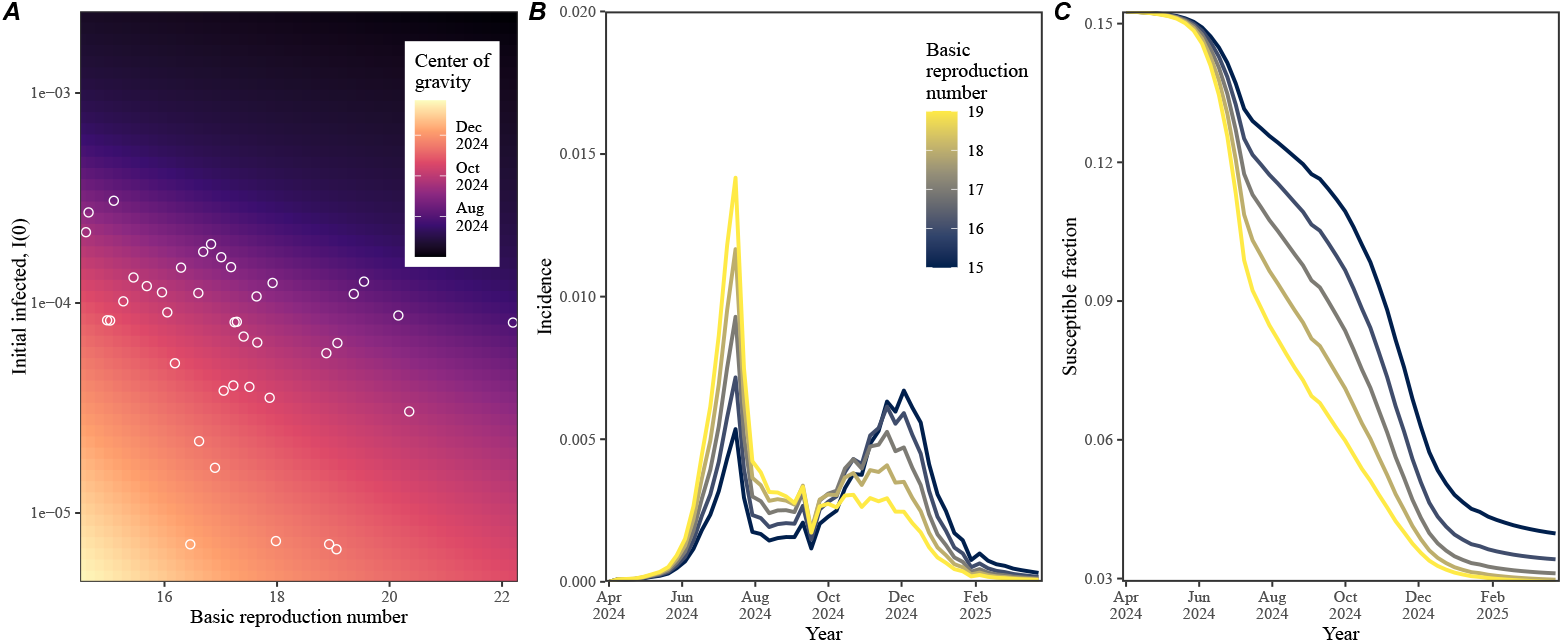
Impact of delayed introduction, contact variation, and susceptible host dynamics on epidemic waves. (A) Relationship between the basic reproduction number ℛ_0_ and infected I(0) fraction with center of gravity. Points represent the estimates for each municipality. (B) Impact of basic reproduction number on pertussis epidemic waves. (C) Impact of basic reproduction number on pertussis susceptible host dynamics. Colors in panels B and C represent values of basic reproduction number.

## Discussion

Understanding mechanisms that drive spatiotemporal variation in epidemic dynamics is a major aim for infectious disease dynamics and control [1, 2, 3, 4, 7, 8, 9]. The large pertussis outbreaks in Korea that occurred between 2024 and 2025 presents a unique perspective on a fine-scale spatiotemporal asynchrony heterogeneity in epidemic dynamics: some municipalities experienced two epidemic waves while other places experienced only the first or second without clear patterns of synchrony. I propose that this variation can be explained by the differences in introduction delays, contact variation, and susceptible host dynamics, where high levels of transmission can cause a large first wave and prevent the second wave through susceptible depletion. Incorporating this idea into a simple transmission model by allowing for a joint estimation of a shared transmission term confirms this hypothesis. Together, these findings shed light into mechanisms that can generate spatial asycnrhony in epidemic dynamics, offering a general framework for understanding the spatiotemporal dynamics of re-emerging infections.

Even though the model presented in this paper explains the observed dynamics reasonably well, there are several limitations to this work. First, the model is purely deterministic and does not account for stochasticity, which is likely an important factor for characterizing the establishment of infections after the initial invasion and the subsequent dynamics, especially in small populations [42, 43, 44]. Second, I estimated that variation in contact rates can explain the observed epidemic patterns, but I was unable to identify the mechanism that explains this variation. As an alternative hypothesis, I also considered the possibility that spatial asycnrhony can be explained by susceptible variation given the role of waning immunity in driving pertussis re-emergence [16, 45, 46]. However, I still did not find a clear correlation between past vaccine coverage and estimated variation in susceptible fraction. Detailed serological data may allow us to test different model formulations and further quantify potential variation in population-level susceptibility [47, 48, 49].

Resulting parameter estimates must be interpreted with care as the model relies on several simplifying assumptions. For example, the main model assumed the same values of initial susceptible fraction across all municipalities and tried to estimate separate basic reproduction number ℛ_0_. Therefore, any regional variation in *S*(0), driven by differences in vaccine coverage and waning, would be captured in the estimate of the ℛ_0_. I also did not consider an age structure in the model, but age-structured contact patterns play an important role in shaping pertussis transmission [33]. For example, [50] recently suggested transmission among elderly may contributed to the persistence of pertussis in Korea. Differences in age structure across regions may also contribute to variation in contact rates and population-level immunity [51], but detailed age structured data would be needed to address these challenges. Despite these limitations, this work provides a quantitative foundation and empirical evidence for understanding epidemic asynchrony.

The continued emergence of new pathogens and re-emergence of vaccine preventable infections, such as pertussis, highlights the importance of identifying hotspots of future outbreaks and developing targeted intervention strategies [52, 53, 48]. The apparent asynchrony in pertussis epidemic in Korea, driven by the differences in introduction timing and contact rates, underlines the importance of detailed pathogen surveillance for early detection and strategic allocation of prevention and control resources. Immunological surveillance platforms will be particularly useful for evaluating potential risk of pathogen re-emergence in the future [47, 48, 49]. A detailed understanding of spatial variation in contact patterns and population-level immunity will be crucial for predicting spatiotemporal dynamics of future outbreaks.

## Acknowledgements

S.W.P. thanks Bryan Grenfell for helpful discussion. S.W.P. was supported by the New Faculty Startup Fund from Seoul National University.

## Author contributions

S.W.P. conceived of the study, conducted the analysis, and wrote the manuscript.

## Competing interests

The authors declare no competing interests.

## Data availability

All data and code used in this paper are available on https://github.com/parklab-snu/asynchrony-pertussis.

## Supplementary Figures

**Figure S1:**
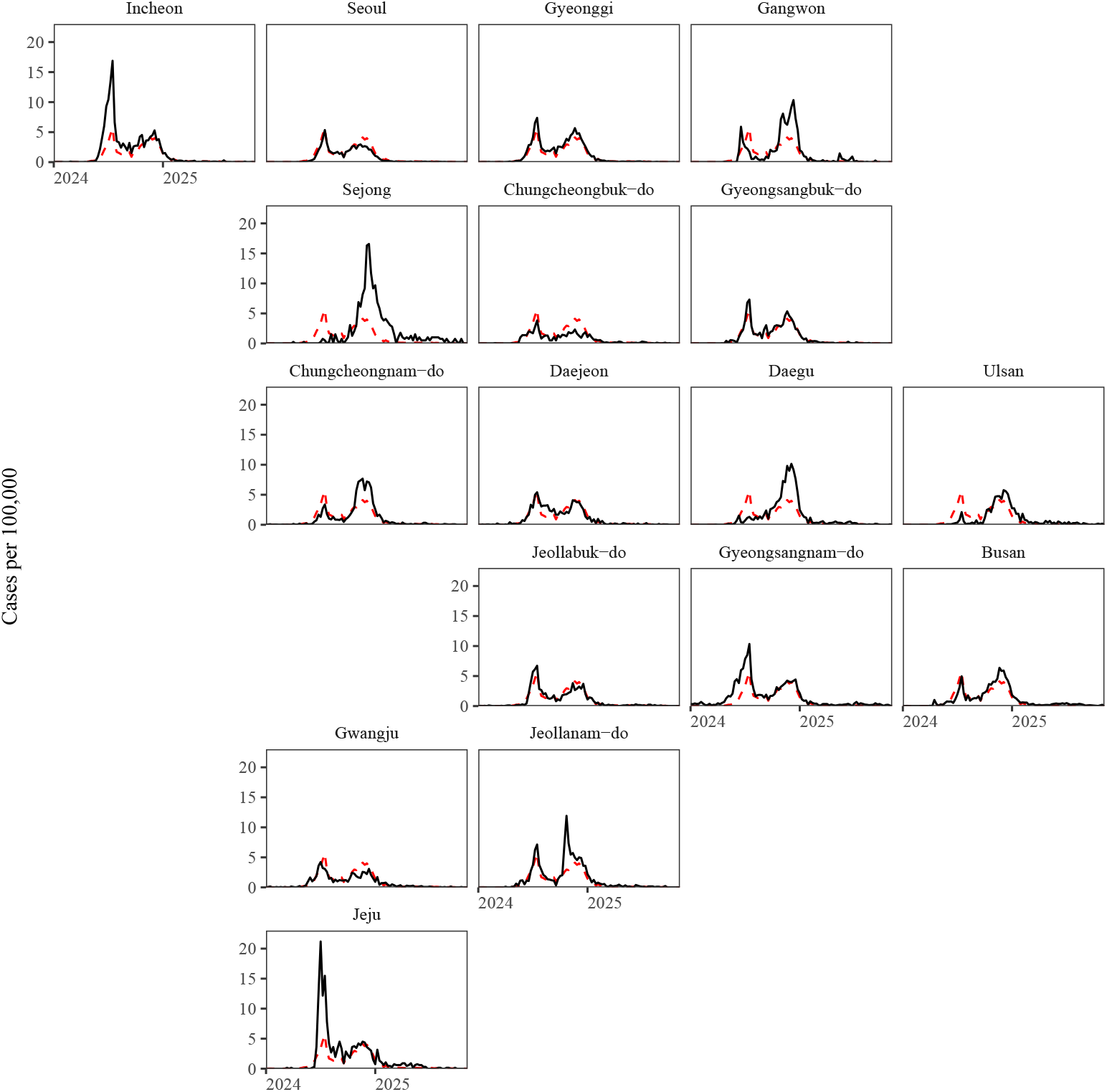
Spatiotempordal dynamics of pertussis outbreak in Korea across first-level administrative divisions, 2024—2025. Time series are arranged based on their approximate locations.

**Figure S2:**
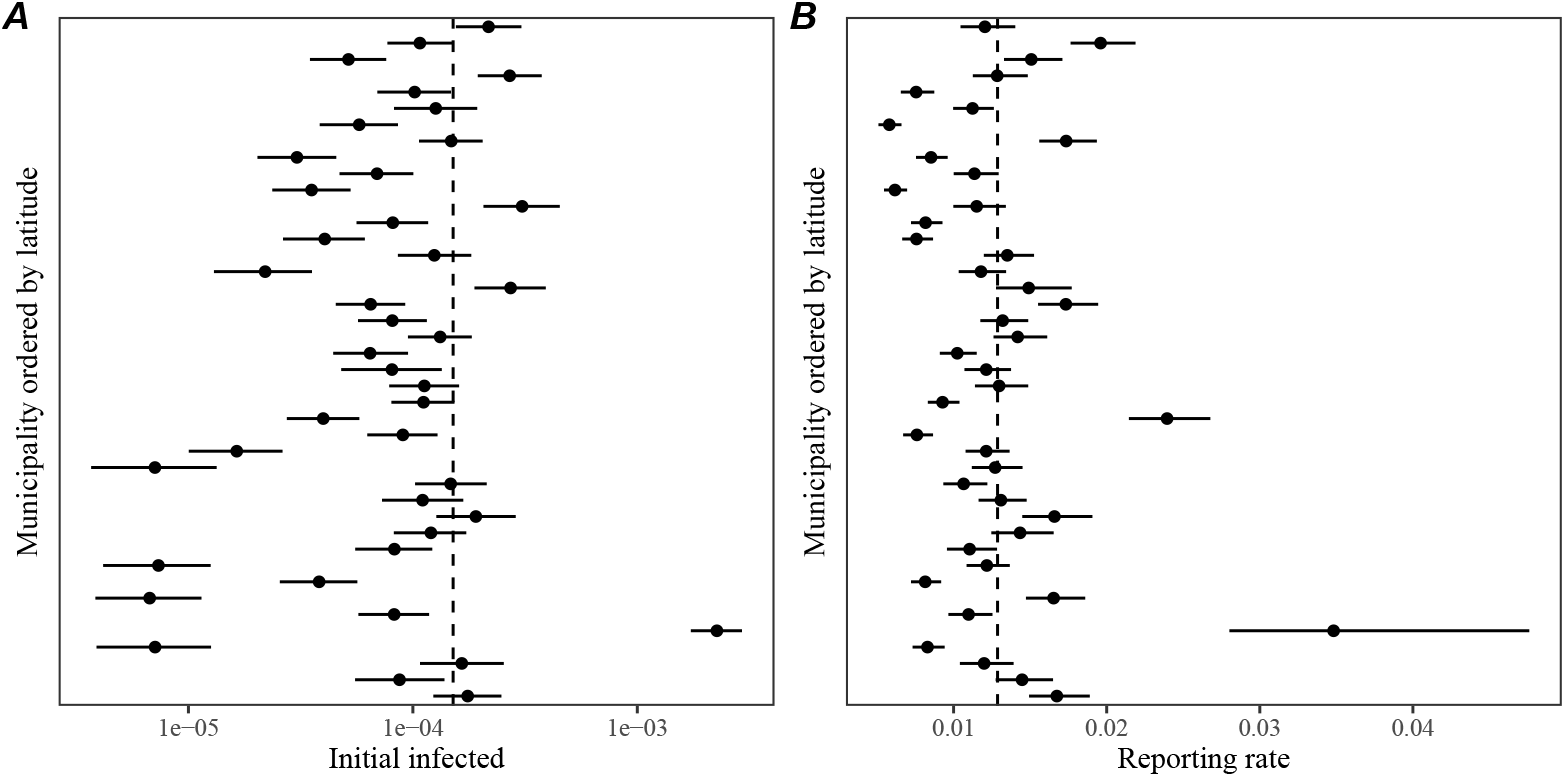
Estimated initial infected fraction (A) and reporting rates (B) using time series data from municipalities with less than 400 total cases. Points represent posterior medians. Error bars represent 95% credible intervals.

**Figure S3:**
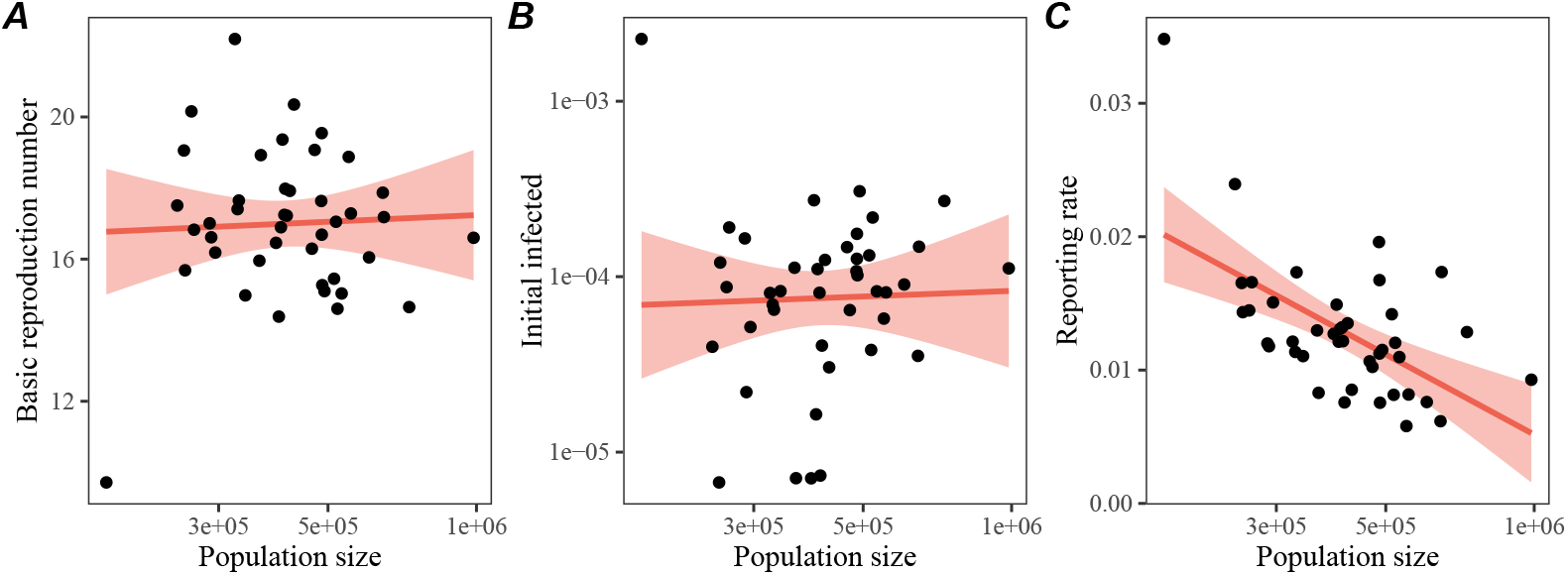
Relationship between population size and parameter estimates for the basic reproduction number (A), initial infected fraction (B), and reporting rate (C). Points represent posterior medians. Error bars represent 95% credible intervals. Red lines and shaded regions represent the linear regression fit and associated 95% confidence intervals.

**Figure S4:**
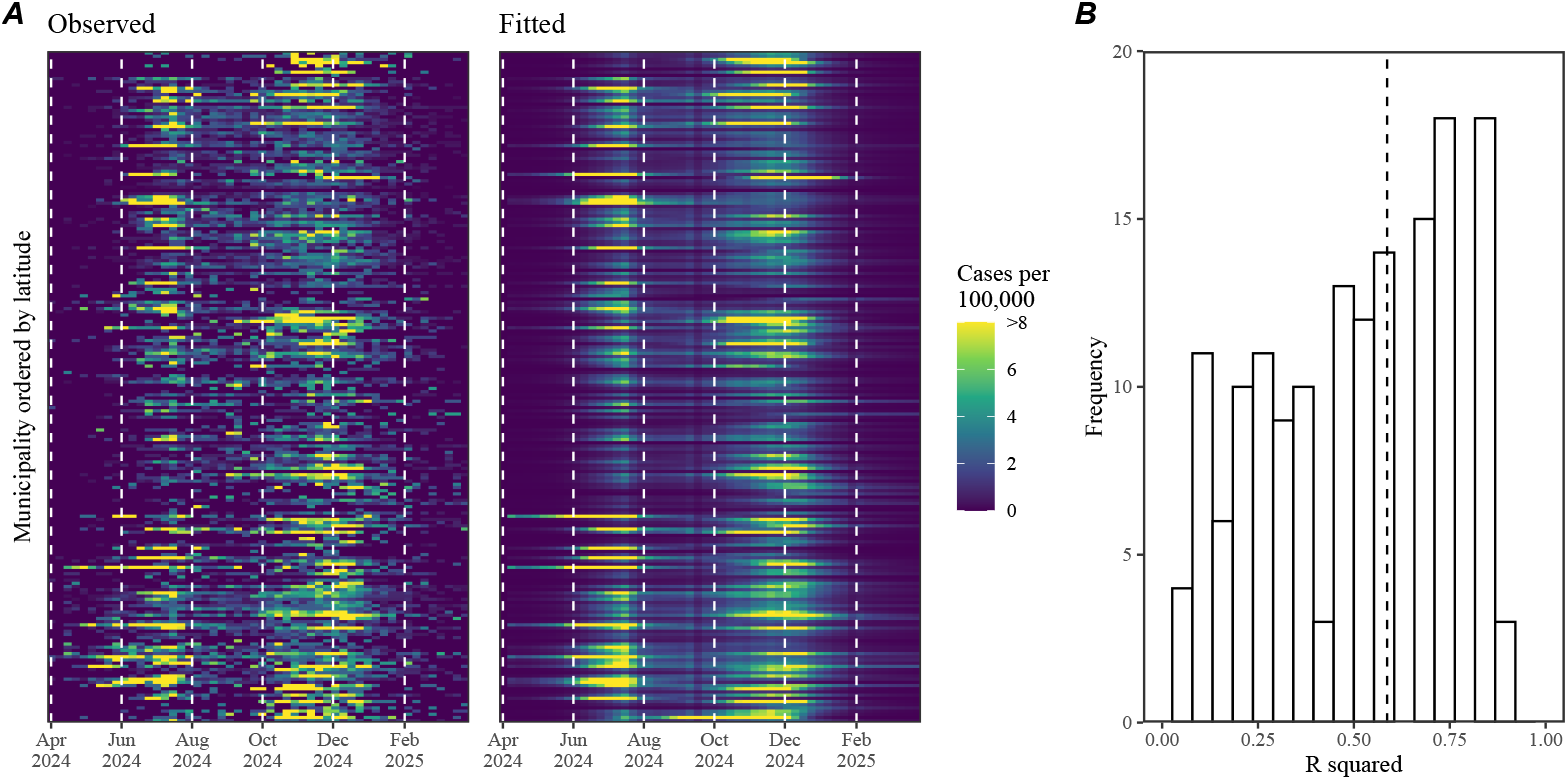
Validation of the transmission model using time series data from municipalities with less than 400 total cases. (A) Comparisons between the observed and predicted epidemic dynamics across municipalities with less than 400 total cases, ordered by latitude. (B) The bar plot represents the distribution of R squared values for model fits. The vertical dashed line represents the median. Initial infected, initial susceptible, basic reproduction number, and reporting probability were estimated for each region while assuming a known transmission variation term *δ*(*t*).

**Figure S5:**
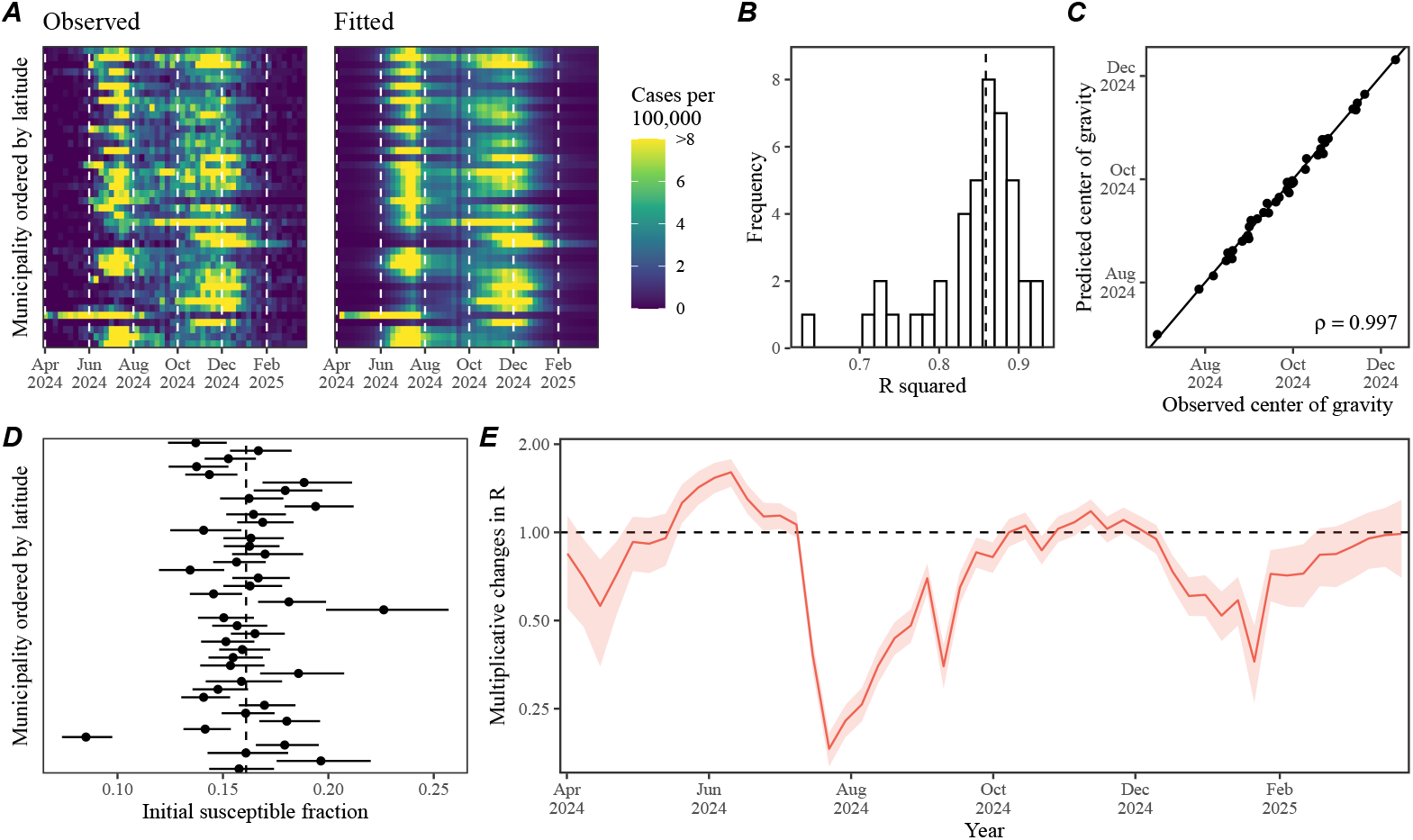
Alternative SEIR model fits assuming variation in initial susceptible fraction. (A) Comparisons between the observed and predicted epidemic dynamics across municipalities with more than 400 total cases, ordered by latitude. (B) The bar plot represents the distribution of R squared values for model fits. The vertical dashed line represents the median. (C) Relationship between the estimated and predicted center of gravity. The solid line indicated the one-to-one relationship. (D) Estimated initial susceptible fraction *S*(0) across municipalities, ordered by latitude. Points and error bars represent posterior medians and corresponding 95% credible intervals. (E) Estimated time-varying pertussis transmission multiplier. The red line and shaded regions represent the posterior median and the corresponding 95% credible interval.

**Figure S6:**
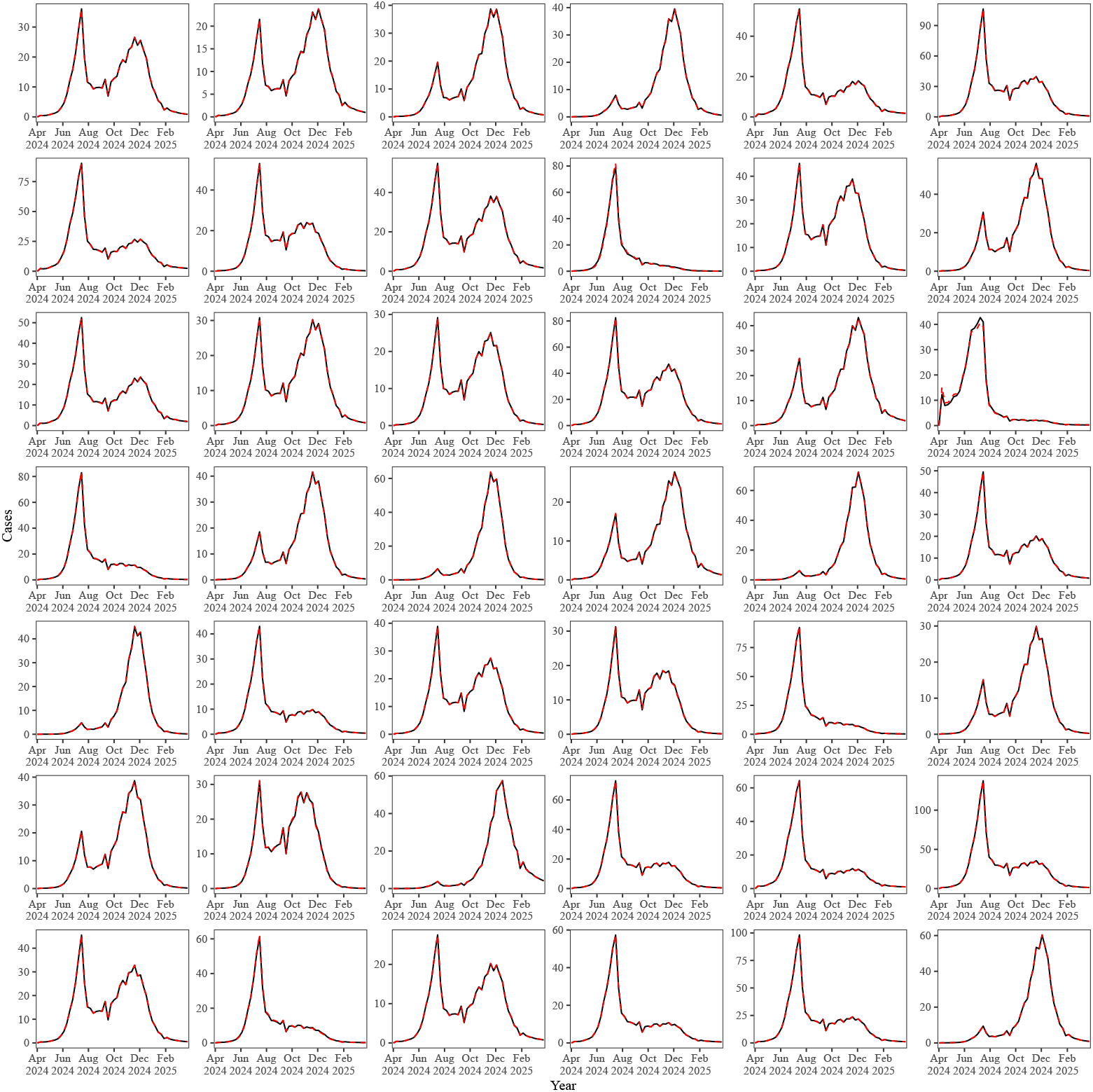
Comparisons between model fits across two different models. Solid lines represent posterior median predictions based on the model assuming separate ℛ_0_ in each region. Red lines represent posterior median predictions based on the model assuming separate *S*(0) in each region.

**Figure S7:**
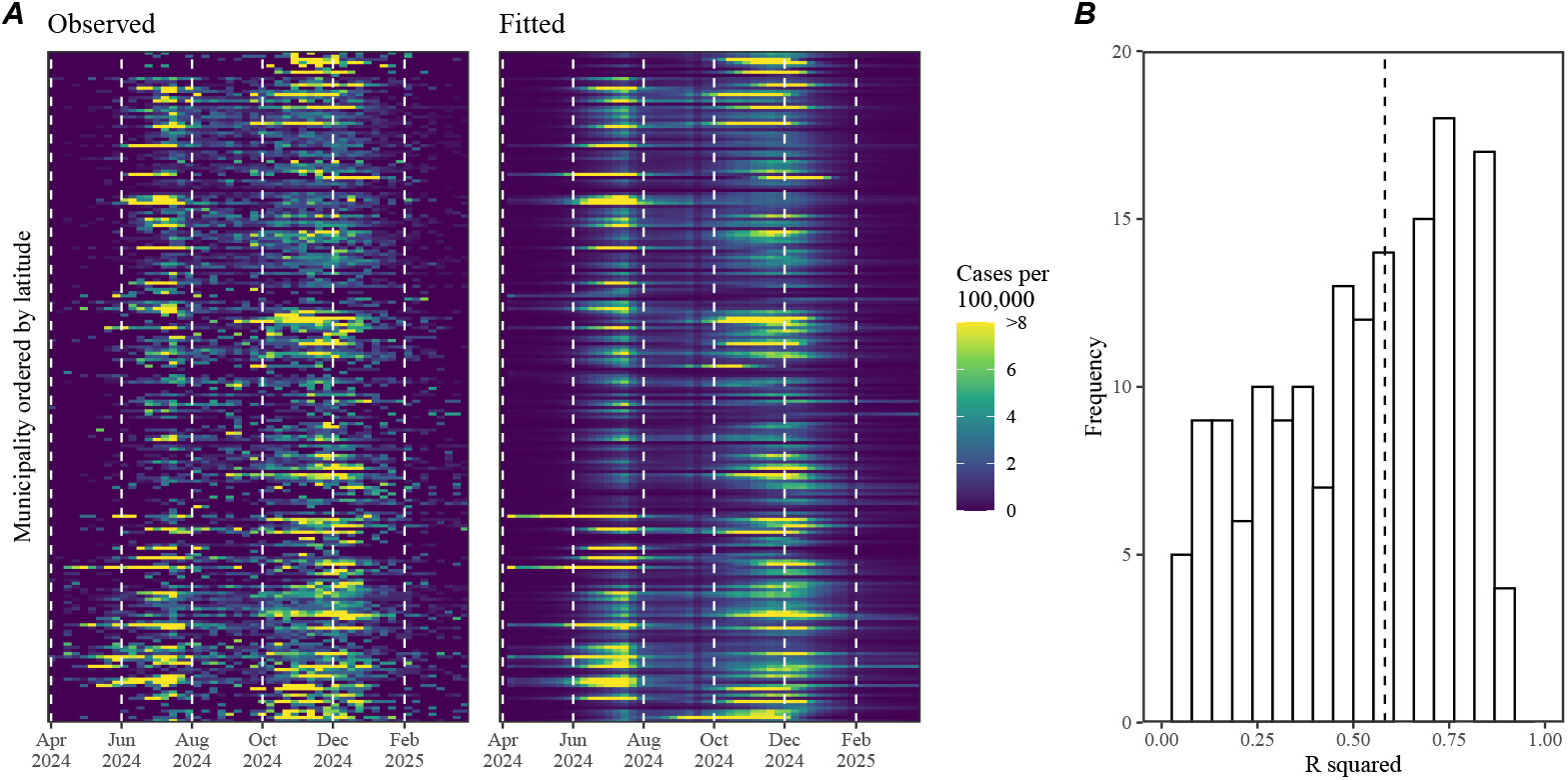
Validation of the alternative transmission model using time series data from municipalities with less than 400 total cases. (A) Comparisons between the observed and predicted epidemic dynamics across municipalities with less than 400 total cases, ordered by latitude. (B) The bar plot represents the distribution of R squared values for model fits. The vertical dashed line represents the median. Initial infected, initial susceptible, and reporting probability were estimated for each region while assuming a known transmission variation term *δ*(*t*) and basic reproduction number.

**Figure S8:**
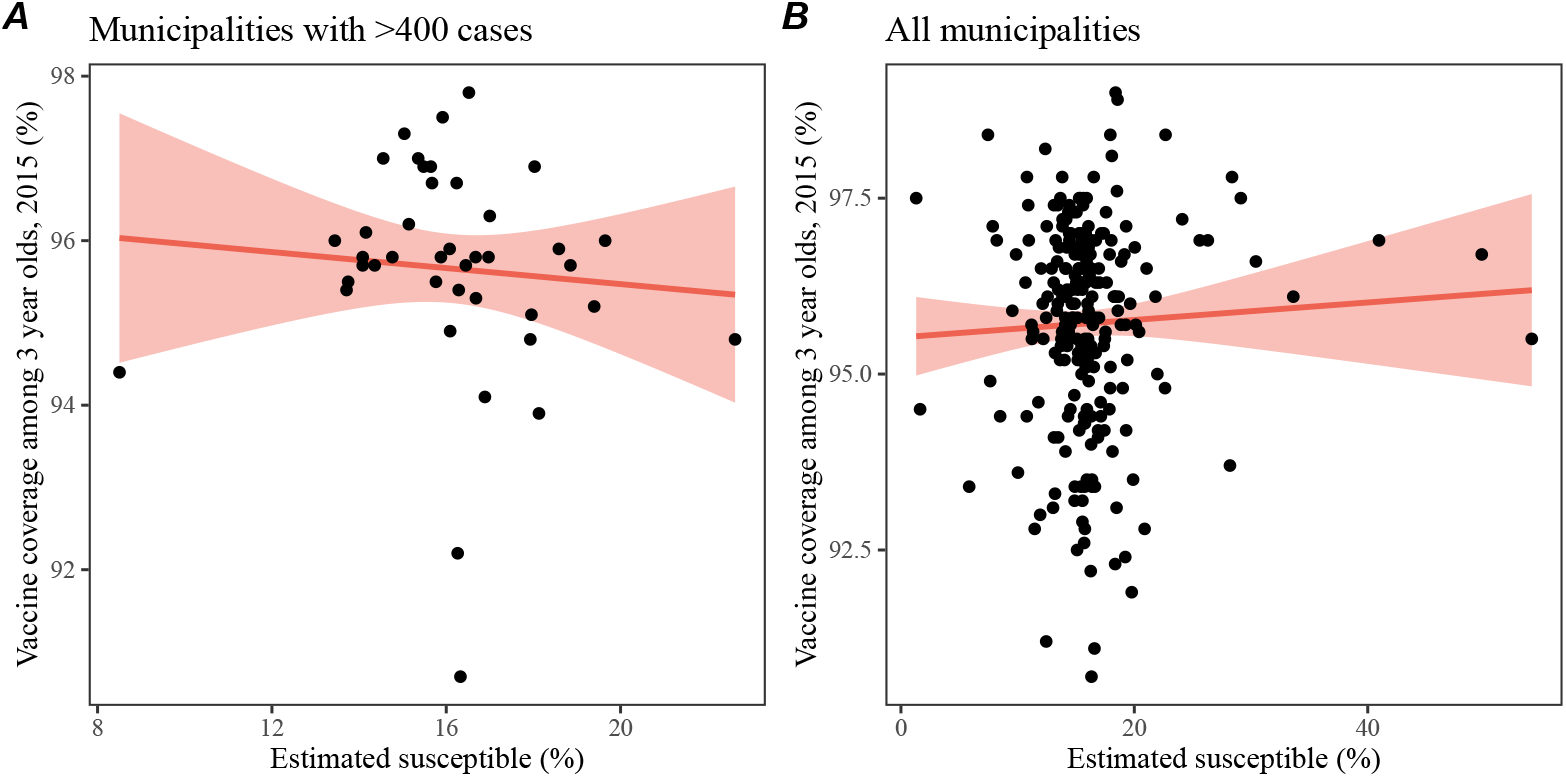
Relationship between estimated initial susceptible and vaccine coverage for (A) municipalities with *>* 400 cases and (B) for all municipalities. Each point represents the estimate initial susceptible and vaccine coverage value for each municipality. Red lines and shaded regions represent the linear regression fit and corresponding 95% confidence intervals.

## Notes

### Competing Interest Statement

The authors have declared no competing interest.

### Summary of Updates

rewrote abstract and fixed typos

